# COVID-19 Test Positivity Rate as a marker for hospital overload

**DOI:** 10.1101/2021.01.26.21249544

**Authors:** Mauro Gaspari

**Affiliations:** Department of Computer Science and Engineering University of Bologna, Italy

**Keywords:** COVID-19, test positivity rate, antigen test, standardization

## Abstract

The use of antigen tests for the diagnosis of COVID-19 in Italy has risen sharply in autumn 2020. Although, Italian regions like Alto Adige, Veneto, Toscana, Lazio, Piemonte and Marche did a large use of these tests for screening and surveillance purposes or for implementing diagnosis protocols, in addition to molecular tests, they were not reported in the statistics in the last months of 2020. As a consequence of this situation the test positivity rate (TPR) index, defined as the number of new positive cases divided by the number of tests, has lost in accuracy. Only in the recent days, starting from the 15th of January 2021, antigen tests have become part of the statistics for all the Italian regions. Despite the lack of data, we have noticed that TPR has a strong correlation with the number of patients admitted in hospitals, and that TPR peaks in general precede the peaks of hospitalized people which occur on average about 15 days later.

In this paper, we have deepened this intuition, analysing the TPR course and its relationship with the number of hospitalized people. To conduct the study we have defined a novel version of the TPR index which takes into account the number of tests done with respect to the population (considering both molecular and antigen tests), the number of infected individuals, and the number of patients healed. Successively, starting from a limited set of data which were made available in November 2020, we have reconstructed the antigen tests time series of four Italian regions, and we computed the TPR index for them.

The results show that TPR peaks precede peaks of hospitalized people in both the first and the second phases of the pandemic in Italy, provided that antigen tests are considered. Moreover, the TPR index trend, can be used to deduct important information on the course of the epidemic, and on the impact of COVID-19 in the health care system, which can be monitored in advance.

## 1 Introduction

In spite of the fact that antigen tests for the diagnosis of COVID-19 have been extensively used in Italy since October 2020, they were not reported in a standardized form in the data provided by the Italian Protezione Civile Department [17] in the last months of 2020. In some Italian regions like Piemonte, Lazio and Marche antigen tests data were mixed with molecular tests, while for regions like Veneto, Toscana and Alto Adige they were partially reported only as additional text comments in the csv file, and for other regions the real situation concerning their use was unknown.

The consequence of this lack of data is that, it was not possible to compute a reliable test positivity rate (TPR) index in the last months of 2020, comparing it in all the 21 regions of Italy.

This inconsistency in the provided data is only a minor drawback, indeed TPR appears to be a powerful tool for monitoring the evolution of the epidemic course that it was not possible to study accurately. Traditionally, TPR has been used to estimate the incidence of diseases in the population, for example for malaria disease [1]. For a similar purpose, it has been recently used to estimate COVID-19 prevalence in the different states of US [10]. It is also recommended by World Health Organization to test positivity proportion from sentinel sites to establish levels of community transmission [11]. However, beyond these well known applications, the large amount of data collected in the COVID-16 pandemic, would allow researcher to explore the use of TPR in different contexts, that previously could not be addressed due to the lack of epidemiological data. For example, we used the TPR combined with epidemiological data in the development of a simulator for reproducing the outbreak process and estimating the impact of the disease in the population at the end of June 2020[18]. The simulator was able to forecast the number of infected people in Italy as confirmed by the serological survey of the Italian Institute for Statistics (ISTAT) published the 3rd of August 2020 [19]. More precisely, the number of case computed by the simulator were in the ranges of the data reported by the ISTAT survey for 16 out of 21 regions, for the whole Italy, and the remaining regions were not far away.

The simulator used the daily growth percentage of official cases to estimate how unknown cases grow adding a variable part which depends on TPR and on a factor *K*. If the number of asymptomatic cases increases like in the summer *K* increases, vice-versa it decreases. The factor *K* is quite difficult to estimate, whilst the TPR can be effectively computed. While tuning the simulator in the second phase of the pandemic an interesting correlation has emerged: the peaks of the daily TPR index were apparently related to the peaks of hospitalized patients in some of the regions. We did an extensive analysis of this marker, showing that peaks of patients in hospitals and intensive care units occurred on average 16 and 12 days after the TPR peaks in the first phase of the pandemic, and that this forecasting capability is preserved in the second phase if data on antigen tests are used.

In this paper, the results of this research are presented. To conduct the study, we have defined an novel version of TPR index that can be computed daily and takes into account the number of tests done considering both molecular and antigen ones, the tests used for patients healed, and the number of infected individuals with respect to the population. Quoting a recent report of the World Health Organizzation: if the TPR is used in a general (non-sentinel) setting *it is heavily influenced by testing strategy and capacity* [11]. As far as we know the TPR has not yet being standardized for COVID-19, our proposal can be considered one of the first attempts to achieve this goal. Although, the weekly index computed in the ISS reports [14] is more accurate than the one we are proposing, it is outdated, namely it estimates the positivity rate more than one-week back with respect to when it is calculated. It cannot be computed daily because many data have to be collected and carefully analysed to discard most of the tests not used for diagnosis purpose.

In our opinion, the results presented in this study could have an enormous importance for the monitoring of the pandemic. What we show is that, if appropriately tuned and standardized the TPR index could become a tool able to predict in advance the impact of the disease in the health care system, to analyse the situation in different areas of the country, and promptly take appropriate actions where this appears necessary.

We conclude the paper, by presenting a proposal for an extended version of the TPR index to be used if data on antigen tests are made available in a correct and more structured form, like it recently happened in Italy. This extended TPR index provides a better integration of antigen tests with molecular tests data, also including vaccinate people.

## 2 The test positivity rate index

Test positivity is one of the metrics commonly used to infer the level of transmission of a disease in a population, it is one of the indexes that the Italian institute for health ISS uses for monitoring the COVID-19 pandemic in Italian regions [13, 14, 15].

We define here a novel version of this index, that can be computed daily. The basic daily test positivity rate Θ is defined as follows:

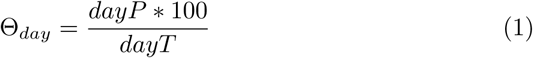

where *dayP* and *dayT* are respectively the new positive cases of the day and the number of tests, e.g. nasopharyngeal swab (NS), done in same day.

To address data bias, indeed data include several anomalies, we considered the average value of *dayP* and *dayT* in the last *µ* days, where *µ* is the incubation period. Using the percentage of positive tests administered over a given period is a common technique to estimate this index [10]. For example, the World Health Organization recommends to compute it from averaged values over a two-week period [11].

Another issue concerns modelling the number of tests done in a given day that are used to diagnose the healed. These tests should not be considered in the total amount of tests; we denote them with *dayR*.

The mean TPR index Θ on *µ* days is defined as follows:

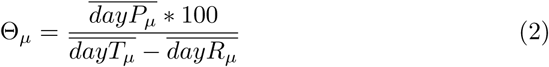

The reasoning behind choosing the length of the incubation period for computing the average values is related to the fact that the set of exposed individuals in a given day will be emptied in about *µ* days, and in the meantime new exposed are added. In other words, if the size of the exposed individuals increases the number of infections detected in the next *µ* days will increase at the same pace.

Finally, a factor Φ is added to Θ to model the impact of the number of tests on the remaining susceptible individuals, which are computed removing the total infected cases *I* from the population *N* of a given region.

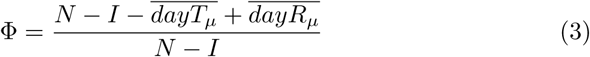

This factor can be also extended to model persons who are vaccinated, removing them like infected individuals. We will consider this issues in the last Section.

The new definition of TPR is the following:

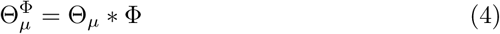

Needless to say, it would be possible to define more precise measures, however this will make the collection of data more difficult. In our opinion our TPR index is a reasonable compromise between what it can be actually collected and effectively represented.

## 3 Correlation with hospitalized patients data

Considering the data provided by the Italian Protezione Civile Deparment [17], we did a detailed analysis to study how the *TPR* index (equation 4) peaks are related to the peaks of hospitalized data.

### 3.1 The first phase

We started from the first phase of the pandemic in February 2020, calculating the *TPR* peaks for each region, and comparing them with hospitalized and intensive unit patients peaks. Hospitalized data includes patients in intensive care units. To address data bias, we discarded the first 7 days at the very beginning of the curve^1^, and then we moved a few days ahead if the TPR is in a descending phase, until it starts increasing again. Starting from this position, we found the highest value of TPR in the first phase, and we computed the distance in days with the hospitalized and intensive unit peaks. The results are reported in Table 1.

**Table 1:** First phase of the pandemic: the first column shows the TPR peak dates and values for all the Italian regions; the second column shows the peak of hospitalized patients, their number and the distance in days from the TPR peak; the third column does the same for patients in intensive units; and the last two columns present the increment percentage of hospitalized patients at the TPR peak to reach the maximum.

| Region | TPR peak<br>Date: rate | Hosp days peak<br>Date:cases,days | IU days peak<br>Date:cases,days | Hosp<br>inc% | UI<br>inc% |
| --- | --- | --- | --- | --- | --- |
| Abruzzo | 21-03:24.36 | 03-04:(437, 13) | 03-04:(76, 13) | 47.6 | 42.11 |
| Basilicata | 18-03:17.76 | 13-04:(76, 26) | 28-03:(19, 10) | 85.53 | 89.47 |
| AltoAdige | 25-03:13.98 | 07-04:(389, 13) | 08-04:(65, 14) | 40.87 | 38.46 |
| Calabria | 16-03:11.33 | 03-04:(200, 18) | 25-03:(23, 9) | 78.5 | 69.57 |
| Campania | 18-03:25.32 | 05-04:(717, 18) | 24-03:(181, 6) | 78.94 | 86.74 |
| Emilia-R. | 06-03:41.61 | 02-04:(4310, 27) | 05-04:(375, 30) | 89.56 | 85.87 |
| Friuli | 19-03:39.17 | 29-03:(296, 10) | 03-04:(61, 15) | 44.93 | 52.46 |
| Lazio | 24-03:14.01 | 27-04:(1607, 34) | 11-04:(203, 18) | 47.67 | 53.69 |
| Liguria | 24-03:41.55 | 31-03:(1332, 7) | 31-03:(179, 7) | 28.68 | 17.88 |
| Lombardia | 20-03:60.78 | 04-04:(13328, 15) | 03-04:(1381, 14) | 34.09 | 23.97 |
| Marche | 17-03:56.85 | 29-03:(1168, 12) | 31-03:(169, 14) | 39.38 | 35.5 |
| Molise | 16-03:26.09 | 01-04:(40, 16) | 27-03:(9, 11) | 80.0 | 44.44 |
| Piemonte | 23-03:42.33 | 07-04:(3985, 15) | 01-04:(453, 9) | 36.34 | 24.28 |
| Puglia | 24-03:15.97 | 04-04:(780, 11) | 05-04:(159, 12) | 52.05 | 64.15 |
| Sardegna | 19-03:19.64 | 05-04:(151, 17) | 08-04:(31, 20) | 65.56 | 70.97 |
| Sicilia | 25-03:15.09 | 06-04:(637, 12) | 25-03:(80, 0) | 46.78 | 0.0 |
| Toscana | 18-03:21.8 | 03-04:(1437, 16) | 01-04:(297, 14) | 59.15 | 46.13 |
| Trentino | 16-03:53.96 | 07-04:(438, 22) | 04-04:(81, 19) | 79.0 | 76.54 |
| Umbria | 19-03:18.75 | 30-03:(220, 11) | 03-04:(48, 15) | 59.09 | 56.25 |
| V.d'Aosta | 31-03:62.33 | 10-04:(147, 10) | 01-04:(27, 1) | 20.41 | 3.7 |
| Veneto | 17-03:12.0 | 01-04:(2068, 15) | 30-03:(356, 13) | 65.23 | 51.97 |

The study shows that the peaks of hospitalized patients occur from about 10 to 18 days after the TPR peak for the majority of regions. The average distance of hospitalized peaks is 16.1 days, and of intensive unit peaks is 12.57 days. The average increment for patients admitted in hospitals was 56.16%, and the average increment for patients in intensive units was 49.25%.

To compute the TPR index, we first assumed an incubation period *µ* of 5 days [20] to average the data. Indeed, we have found that, if the number of considered days deviates from the length of the incubation period, the average difference between TPR peaks and hospitalized patients peaks decreases. Thus, the forecasting capabilities of TPR deteriorates. Table 2 illustrates this result comparing 4 days, 5 days, 6 days, 7 days and 14 days based averages, using the basic version of the index (equation 4): choosing *µ* for computing average values was correct. However, we observed that using 5 days only, a few peaks in some regions still were caused by bias effects on data. So, we have preferred 6 days to average the data, as a compromise to reduce the bias effects maintaining a good forecasting ability. Anyhow, the COVID-19 incubation period ranges on average between 5 and 6 days.

**Table 2:** How different means over N days influence the TPR peak date: on average the peak is pushed forward if the number of considered days deviates from the incubation period.

| Curve | 4 days | 5 days | 6 days | 7 days | 14 days |
| --- | --- | --- | --- | --- | --- |
| Hosp | 15.9 | 16.9 | 16.1 | 14.57 | 13.38 |
| IU | 12.38 | 13.8 | 12.57 | 11.05 | 9.86 |

A critical point of the factor Φ (equation 3) is that it needs the number of infected people to be computed, but real cases are actually many more then those reported in official data, as pointed out in several studies, for example [21, 12, 18]. This was confirmed for Italy by the Italian Statistical Institute serological survey [19]. Given that the simulator we have developed [18] provides a good approximation of unknown cases in the first phase of the Pandemic in Italy, we repeated the study using the number of infected people computed by the simulator for each day. The results show that there are no significant differences and the average distance of peaks remain the same, thus the number of infected people reported in official data can be actually used for computing the TPR index in the first phase of a pandemic.

Looking back to Table 1, we note that for some of the regions like, Basilicata, Emilia Romagna, Lazio and Trentino the time to reach the peak is considerably longer than the mean number of 16 days. Observing their graphs one can see that all of them were characterized by an uncontrolled period where the TPR remains high: four weeks for Emilia Romagna (Figure 1) and two weeks for Trentino (Figure 2) where the TPR was about to 35%. For both these regions the percentage increment to reach the peak of hospitalized and IU patients is fairly high (more then 79%). Considering Basilicata (Figure 3) there where one week where the TPR was about to 15%, and the increment of patients rate more than 85%. On the contrary, considering the Lazio region (Figure 4), where the TPR was about to 12%, the time to reach the peak was shorter and the added percentage to hospitalized people not so great (about 47%). As a further example of the first phase it is worth to report the diagram of region Lombardia (Figure 5). Note that unlike other regions the TPR index at the beginning of June 2020 was still significantly above 0. This is one of the motivations of the high number of active unknown cases estimated by our simulator at the beginning of June 2020 for the Lombardia region [18]. The diagrams of the remaining 16 Italian regions are sketched in Figure 6 and 7.

**Figure 1:**
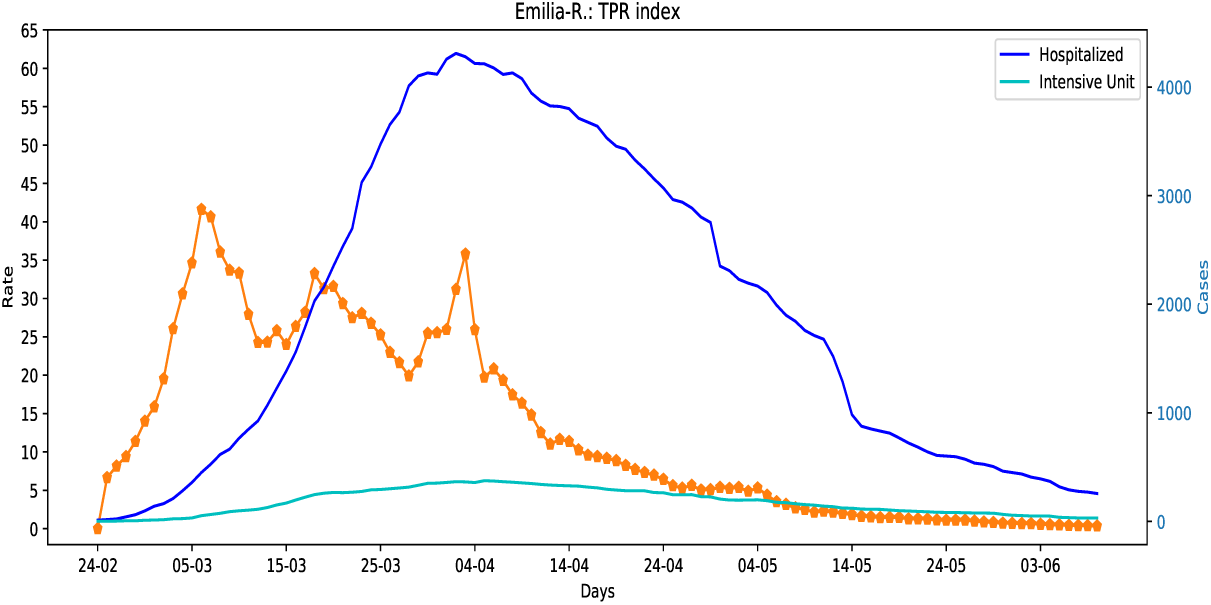
Emilia Romagna region: TPR index compared with hospitalized data.

**Figure 2:**
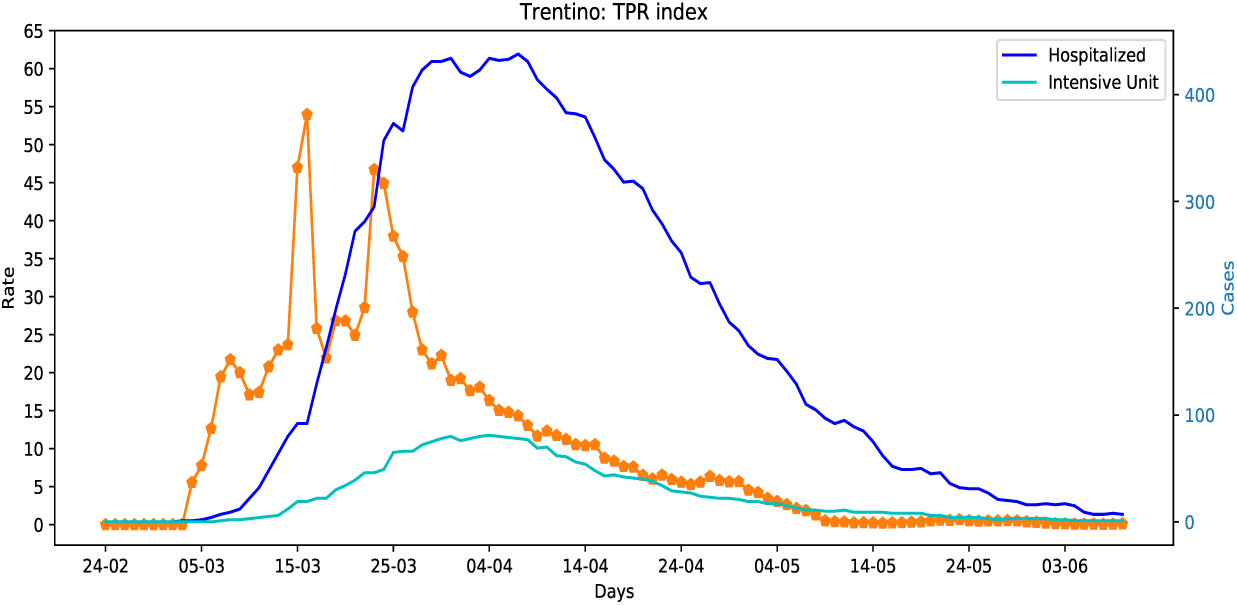
Trentino region: TPR index compared with hospitalized data.

**Figure 3:**
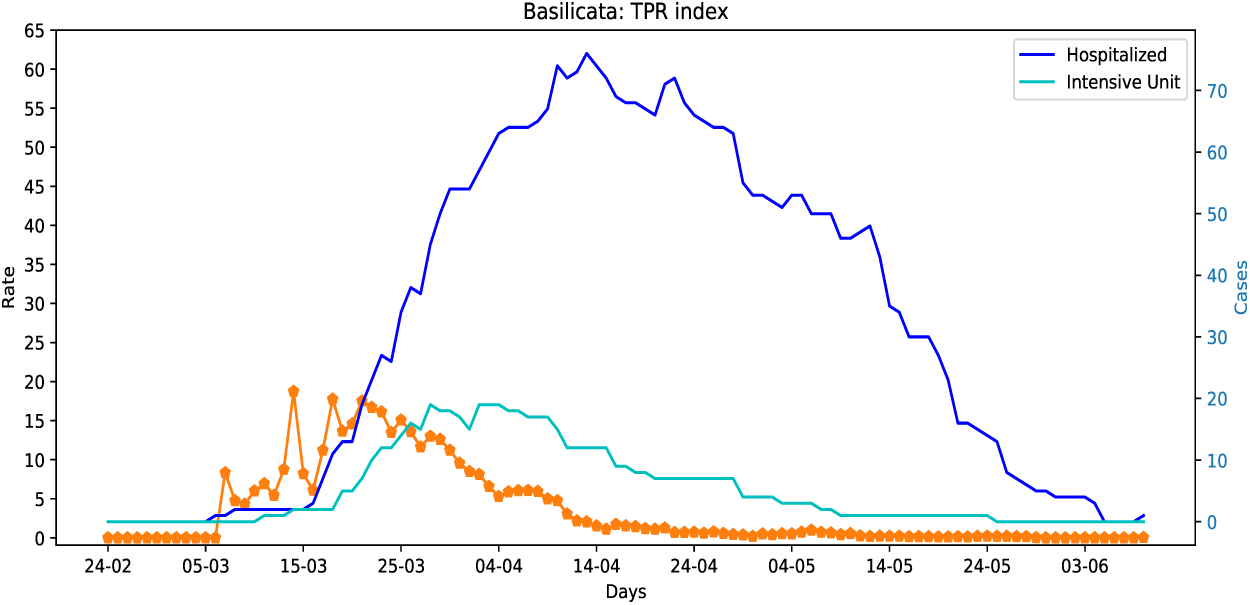
Basilicata region: TPR index compared with hospitalized data.

**Figure 4:**
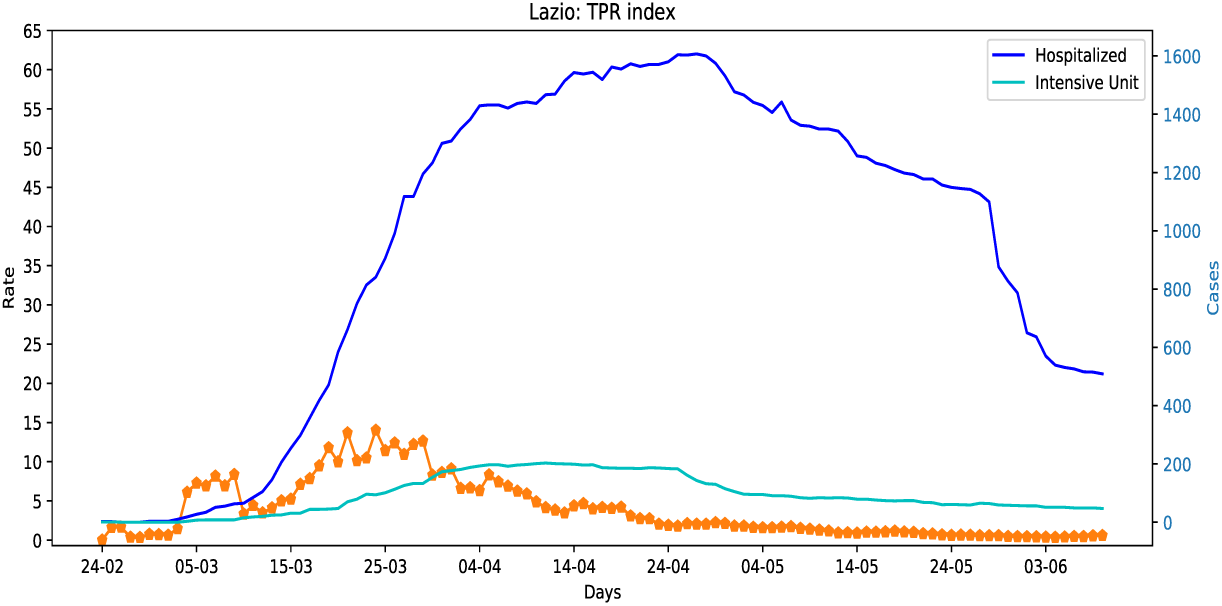
Lazio region: TPR index compared with hospitalized data.

**Figure 5:**
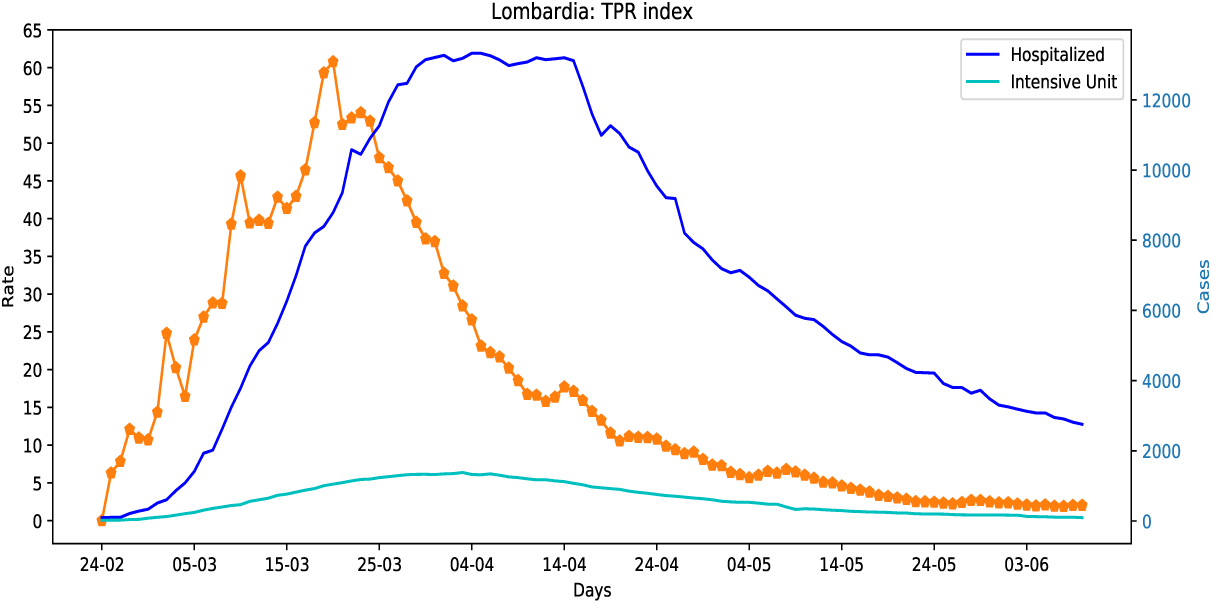
Lombardia region: the TPR index was still above 0 at the beginning of June 2020.

**Figure 6:**
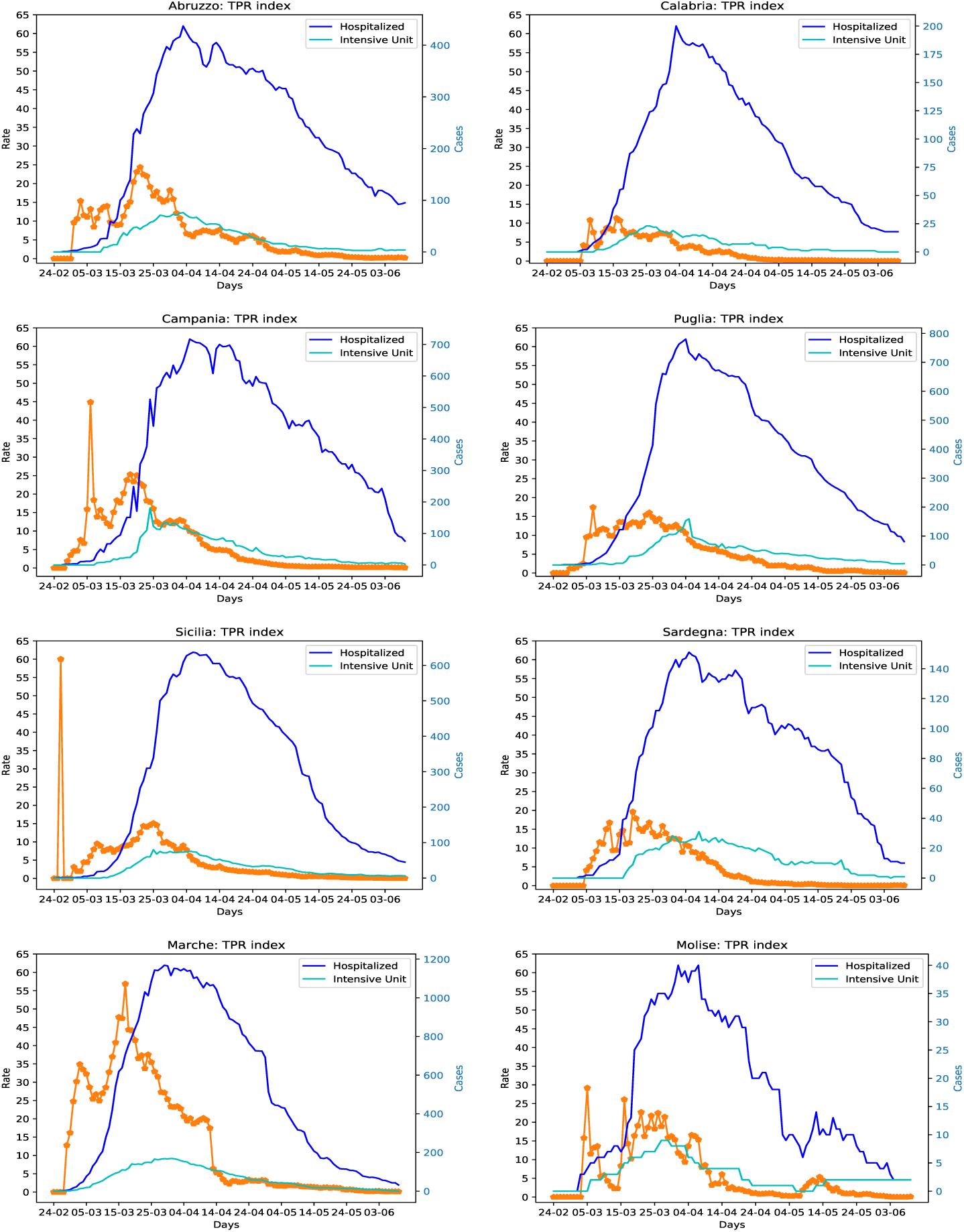
TPR index compared with hospitalized data for regions in the south of Italy and islands.

**Figure 7:**
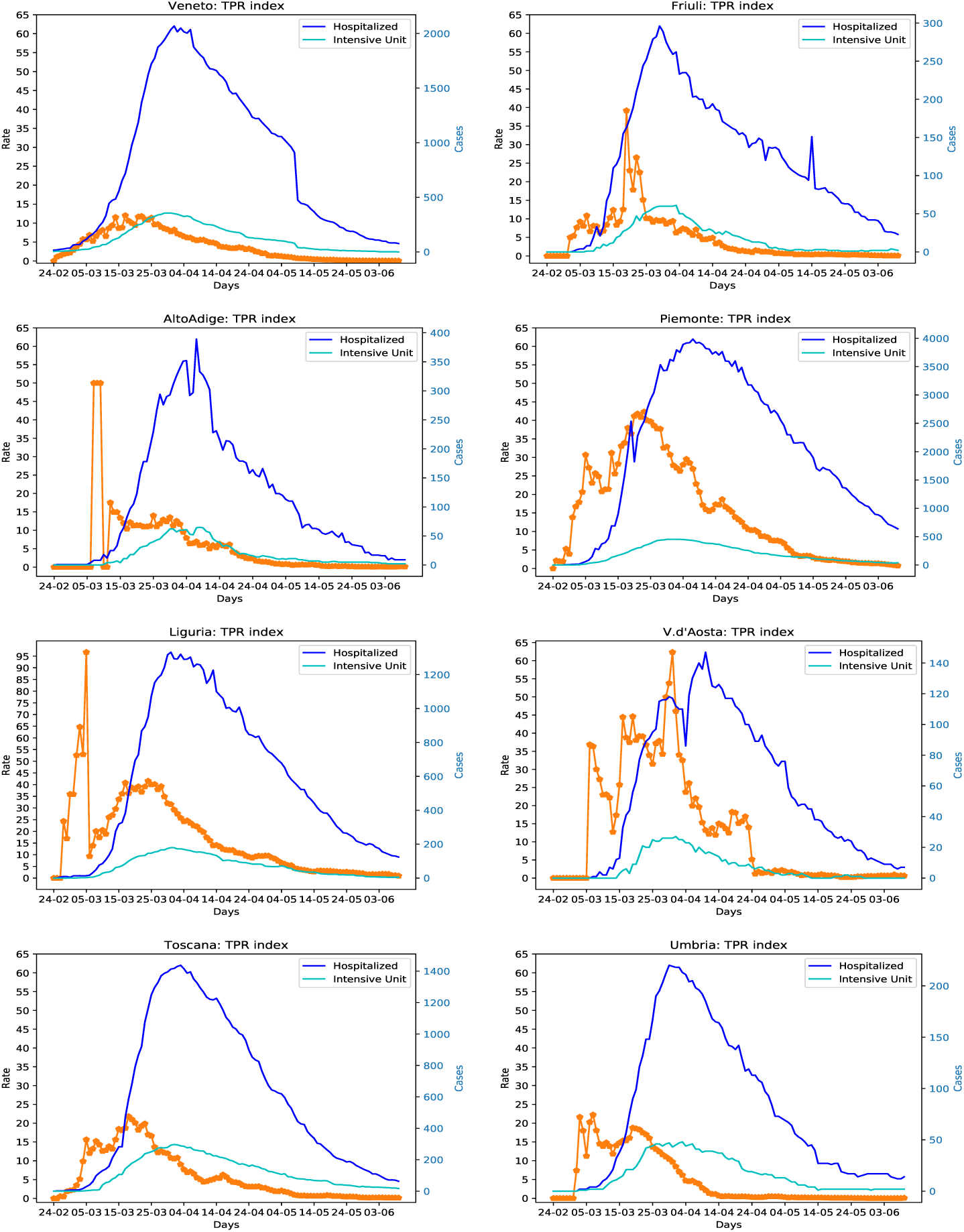
TPR index compared with hospitalized data for regions in the north and center of Italy.

### 3.2 The second phase

The second phase of the epidemic in Italy broke out in autumn. From the perspective of diagnosis tools the second phase is characterized by the growing use of antigen tests [16]. We repeated the study in the second phase (until the 5th of December 2020), starting from the 1st of October 2020 using equation 4 again. In this case the correction of initial bias was not necessary. The 5th of December was a good date for evaluating the TPR index because most of the regions had already reached the peaks of hospitalized persons.

What we got is that the forecasting capabilities of TPR have deteriorated of more than one half in the second phase, obtaining an average of 6.95 and 6.48 days ahead with respect to hospitalized and intensive units peaks respectively. On the contrary the increment of hospitalized people was lower on average: 12.08% for patients admitted in hospitals people and 17.25% for patients in intensive units. Table 3 shows the details for each region.

**Table 3:** Second phase of the pandemic until the 5th of December 2020: the first column shows the TPR peak dates and values for all Italian regions; the second column shows the peak of hospitalized patients, their number and the distance in days from the TPR peak; the third column does the same for patients in intensive units; and the last two columns present the increment percentage of hospitalized patients at the TPR peak to reach the maximum.

| Region | TPR peak<br>Date: rate | Hosp days peak<br>Date:cases, days | IU days peak<br>date:cases, days | Hosp<br>inc% | UI<br>inc% |
| --- | --- | --- | --- | --- | --- |
| Abruzzo | 14-11:17.11 | 30-11:(790, 16) | 28-11:(77, 14) | 23.54 | 23.38 |
| Basilicata | 11-11:14.93 | 04-12:(185, 23) | 17-11:(30, 6) | 17.84 | 36.67 |
| AltoAdige | 13-11:26.92 | 16-11:(530, 3) | 14-11:(44, 1) | 6.79 | 6.82 |
| Calabria | 23-11:17.26 | 23-11:(482, 0) | 17-11:(53, -6) | 0.0 | 11.32 |
| Campania | 07-11:20.16 | 23-11:(2532, 16) | 03-11:(227, -4) | 23.58 | 21.15 |
| Emilia-R. | 26-11:15.55 | 25-11:(3012, -1) | 26-11:(258, 0) | 2.49 | 0.0 |
| Friuli | 23-11:14.5 | 02-12:(710, 9) | 01-12:(62, 8) | 12.68 | 11.29 |
| Lazio | 18-11:10.93 | 27-11:(3762, 9) | 03-12:(364, 15) | 10.31 | 12.64 |
| Liguria | 07-11:20.91 | 14-11:(1510, 7) | 25-11:(123, 18) | 8.61 | 37.4 |
| Lombardia | 19-11:27.77 | 22-11:(9340, 3) | 22-11:(949, 3) | 1.43 | 3.58 |
| Marche | 14-11:20.22 | 29-11:(683, 15) | 25-11:(94, 11) | 18.3 | 21.28 |
| Molise | 21-11:13.96 | 26-11:(78, 5) | 30-11:(14, 9) | 23.08 | 35.71 |
| Piemonte | 11-11:25.54 | 20-11:(5618, 9) | 24-11:(404, 13) | 8.69 | 15.84 |
| Puglia | 05-12:19.31 | 01-12:(1911, -4) | 04-12:(227, -1) | 6.02 | 3.08 |
| Sardegna | 21-11:13.77 | 05-12:(674, 14) | 25-11:(76, 4) | 14.84 | 7.89 |
| Sicilia | 18-11:18.98 | 23-11:(1847, 5) | 26-11:(253, 8) | 4.28 | 5.14 |
| Toscana | 11-11:16.3 | 23-11:(2128, 12) | 22-11:(298, 11) | 11.94 | 17.45 |
| Trentino | 10-11:11.41 | 24-11:(478, 14) | 05-12:(48, 25) | 33.47 | 41.67 |
| Umbria | 09-11:16.68 | 23-11:(449, 14) | 23-11:(78, 14) | 5.57 | 17.95 |
| V.d'Aosta | 19-11:33.42 | 03-11:(183, -16) | 11-11:(17, -8) | 14.75 | 29.41 |
| Veneto | 05-12:28.91 | 28-11:(2963, -7) | 30-11:(320, -5) | 5.47 | 2.5 |

It seems that the TPR index has lost most its forecasting capabilities in the second phase. One possible hypothesis is that the results of the first study have been influenced by the lockdown strategy started the 9th of March 2020.

However, analysing the details of the single regions, it appears that for some of them the TPR forecasting capabilities are preserved: Abruzzo, Basilicata, Friuli, Marche, Toscana, Trentino, Umbria. Nevertheless, for many other a strange effect leaps to the eye: the peaks hospitalized people precede the peak of infections (TPR index), among them Veneto and Emilia Romagna. An explanation of this phenomenon could be that in those regions infections are detected much after, compared to when they occur. Is the pandemic not well controlled in these regions? This is an obvious contradiction, indeed Veneto and Emilia Romagna have probably among the best contact tracing systems in Italy, as illustrated by the ISS weekly reports (see for example [14] and [15]). So a second hypothesis seems to be more plausible: the computed TPR index does not work properly in these cases, because antigen tests which are largely used in Veneto and in Emilia Romagna are not included so far in the official data provided by the Italian Protezione Civile web site [17]. The situation of Veneto is confirmed by the ISS weekly report N.29 [13] and by the notes transmitted by the Veneto region in November 2020 available in the csv (Comma Separated Values) file of italian regions in the Protezione Civile web site [17], in which the antigen tests numbers were included for a short period in the text notes only.

This point was controversial, for regions like Veneto, Toscana and Alto Adige some data on antigen tests were available in the text notes of the csv file [17] in November 2020, but unfortunately the transmission of antigen data was stopped even in the text notes in December 2020. Thus, it was not possible for us to study the last period of the second phase.

Despite this, we were able to collect the data on antigen tests for the above three regions until the 5th of December, extracting them from the csv file notes, when they were available, or searching them on the web and on the news. These data were sufficient for us to evaluate the impact of antigen tests in the TPR index.

In order to perform the new study a TPR index extended with antigen tests should be defined properly.

## 4 Extending TPR with antigen tests data

First we define a basic method that would allow us to compute the TPR index if only the total number of antigen tests done in each day is given. This is actually the situation of data on antigen tests we have collected.

A problem concerning the representation of antigen tests information is that in the considered period almost all the positive ones had to be confirmed by molecular tests. Thus tests of positive cases individuated with antigen tests are repeated two times, and should not be considered in the estimation of TPR. If the number of positive antigen tests would be available, it should be subtracted to the total number of tests to compute the TPR, but unfortunately this is not the case of the data available in the last months of 2020. Additionally, there is a further representation problem concerning repeated tests, because the molecular tests that are needed to confirm the diagnosis could be done in the successive days. We have solved this problem using average values on *µ* days for both molecular and antigen tests, estimating an average number of repeated tests.

In spite of this problem, we can estimate the number *Pr* of repeated tests as follows. Let *P*^*T*^ and *P*^*A*^ be the positivity percentage of molecular and antigen tests, the antigen positivity rate is:

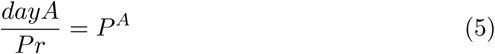

and the molecular positivity rate is:

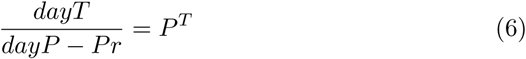

where *dayA* is the number of antigen tests and *dayT* the number of molecular tests in a given day. If we assume that the positivity rates for antigen tests and molecular tests are the same, e.g. *P*^*T*^ =*P*^*A*^, we can estimate the number *Pr* of repeated tests equalizing the two formulas. The number of repeated tests *Pr* can be computed as follows using average values:

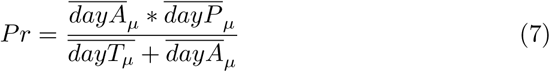

Given that in general *P*^*T*^ *>P*^*A*^, see for example the results presented here [7], the computed number of repeated tests *Pr* can be considered a reasonable upper bound for the estimation. However, if more data will be made available to estimate the difference between the two percentages *P*^*T*^ =*Ψ*\**P*^*A*^, a more accurate definition of *Pr* would be possible:

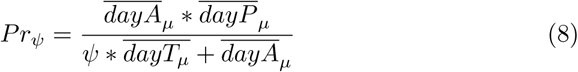

In the following, we will use the upper bound defined in equation 7. The TPR 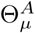 that include antigen tests is defined removing from the total number of tests the repeated ones:

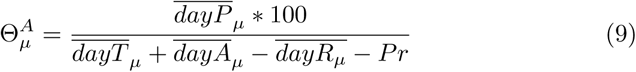

The factor Φ is extended removing antigen tests except duplicates.

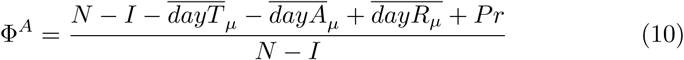

Finally, the TPR index 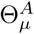 including antigen tests is defined as follows:

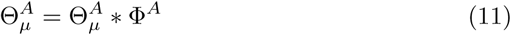

### 4.1 The effect of antigen tests in the TPR index

Considering this extended version of the TPR index, we have analyzed the data of Veneto, Alto Adige and Toscana up to the 5th of December 2020. When available, we used the data that appeared in the csv file notes in the official web site [17], and we have completed them for each region adding an antigen tests column as follows:

- *Toscana*: cumulative antigen tests data where available in the csv file notes [17] from the 22th of October 2020, but for several days the notes were missing. However, for the missing days it was possible for us to get the exact numbers of antigen tests done from the news [25], indeed the regione Toscana reports published daily. We double checked these data with those available in the csv file notes, and we were able to reconstruct the entire series until the 4th of January 2021.
- *Veneto*: cumulative antigen tests data where available in the csv file notes [17] from the 11th of November 2020, in that day the total of 492456 was reported and the other cumulative values were successively added, until the 2nd of December 2020, then disappeared. The values for the successive days has been taken listening to press conferences of regine Veneto. Cumulative values prior to the 10th of November were gradually distributed until the 3rd of October 2020.
- *Alto Adige*: the daily antigen test data were available in the csv file notes [17] since the 6th until the 20th of November 2020, meanwhile a screening campaign has started, involving a large percentage of the population [22, 23]. However, the data of the screening were only partially integrated with those in the official Web site. The screening web site [22] reported the number of antigen tests done in each day and the number positive tests, but only the positive tests were included in the Protezione Civile csv file as notes^2^, not including them among the official positives. Thus, for the period from 21th November to the 24th (the data in the Protezione Civile are reported one day after with respect to the screening ones), we merged screening data with those in the official web site, inserting antigen tests in the new column, and adding the positive cases to official positives until the 27th of November. For the successive days data on antigen tests have been made available in the reports of the Provincia di Bolzano web site [8]. In summary, we were able to rebuild the complete series for Alto Adige integrating screening data.

The results of the study are presented in Table 4, we used equation 11 to compute the TPR index. The table shows that the forecasting capabilities of the TPR index have been restored for the analyzed regions. The average distance of the TPR peaks from the hospitalised people peaks moves from 2 to 12.66 days for these regions, and the average distance for all the Italian regions reaches 8.38 days. The average increment of hospitalize people for the 3 considered regions increases to 22.66% and that of patients in intensive units moves to 20.65%. Figures 8, 9 and 10 illustrate the sliding of the TPR curve using antigen tests in the estimation in Veneto, Toscana and Alto Adige.

**Table 4:** This table presents the results of the study for the regions for which antigen test data were available in November 2020, the organization of data is the same of the previous Tables.

| Region | TPR peak<br>Date: rate | Hosp days peak<br>Date:cases,days | IU days peak<br>date:cases,days | Hosp<br>inc% | UI<br>inc% |
| --- | --- | --- | --- | --- | --- |
| AltoAdige | 09-11:20.8 | 16-11:(530, 7) | 14-11:(44, 5) | 14.72 | 6.82 |
| Toscana | 04-11:14.81 | 23-11:(2128, 19) | 22-11:(298, 18) | 28.76 | 33.89 |
| Veneto | 16-11:9.47 | 28-11:(2963, 12) | 30-11:(320, 14) | 24.5 | 21.25 |

**Figure 8:**
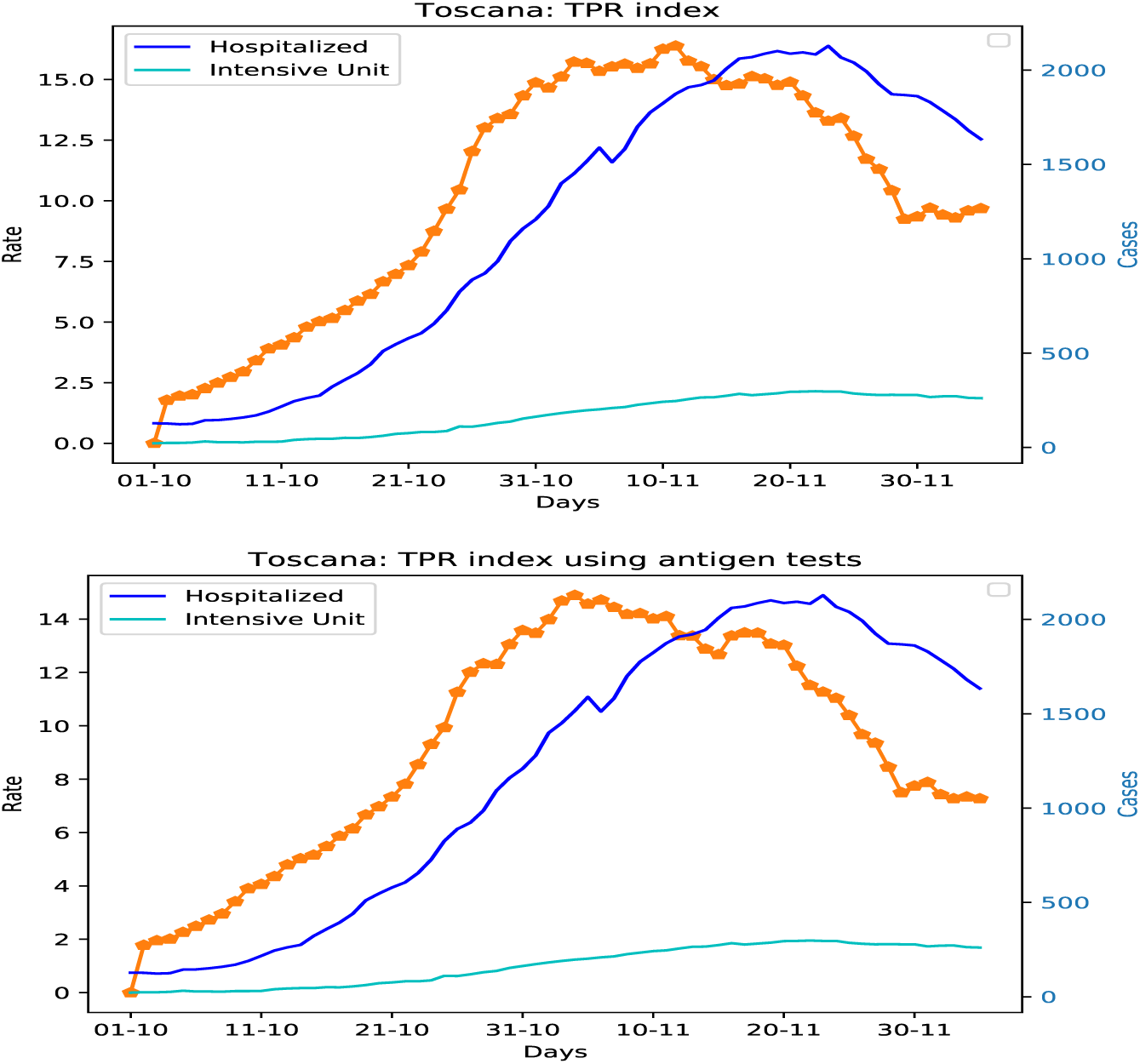
Toscana region: TPR index sliding compared with hospitalized data. The diagram in the bottom includes antigen tests in the estimation.

**Figure 9:**
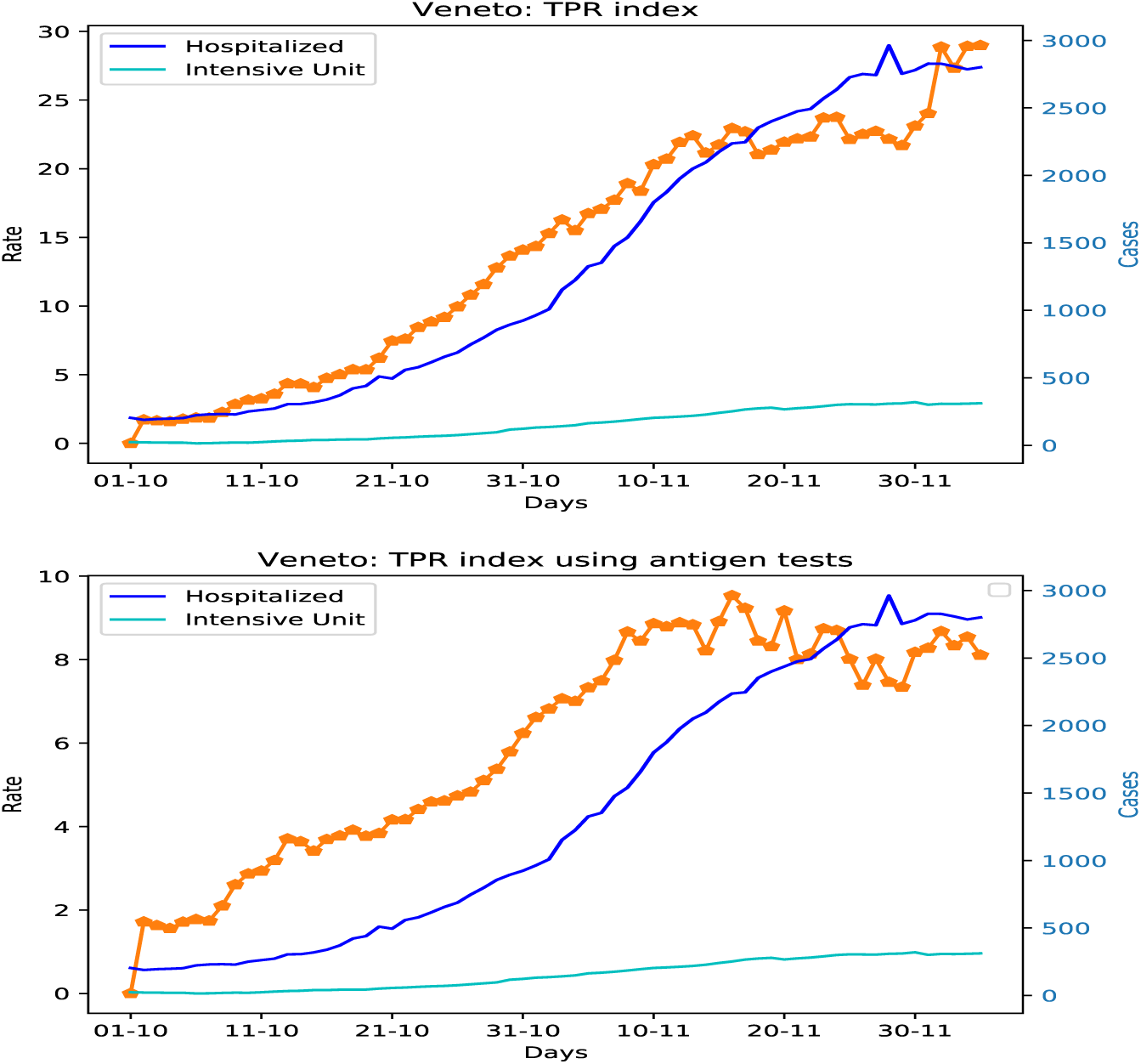
Veneto region: TPR index sliding compared with hospitalized data. The diagram in the bottom includes antigen tests in the estimation. The daily data on antigen test before the 11th of November 2020 were not available, we distributed the total number of tests reported for the 11th of November in the previous days until the beginning of October.

**Figure 10:**
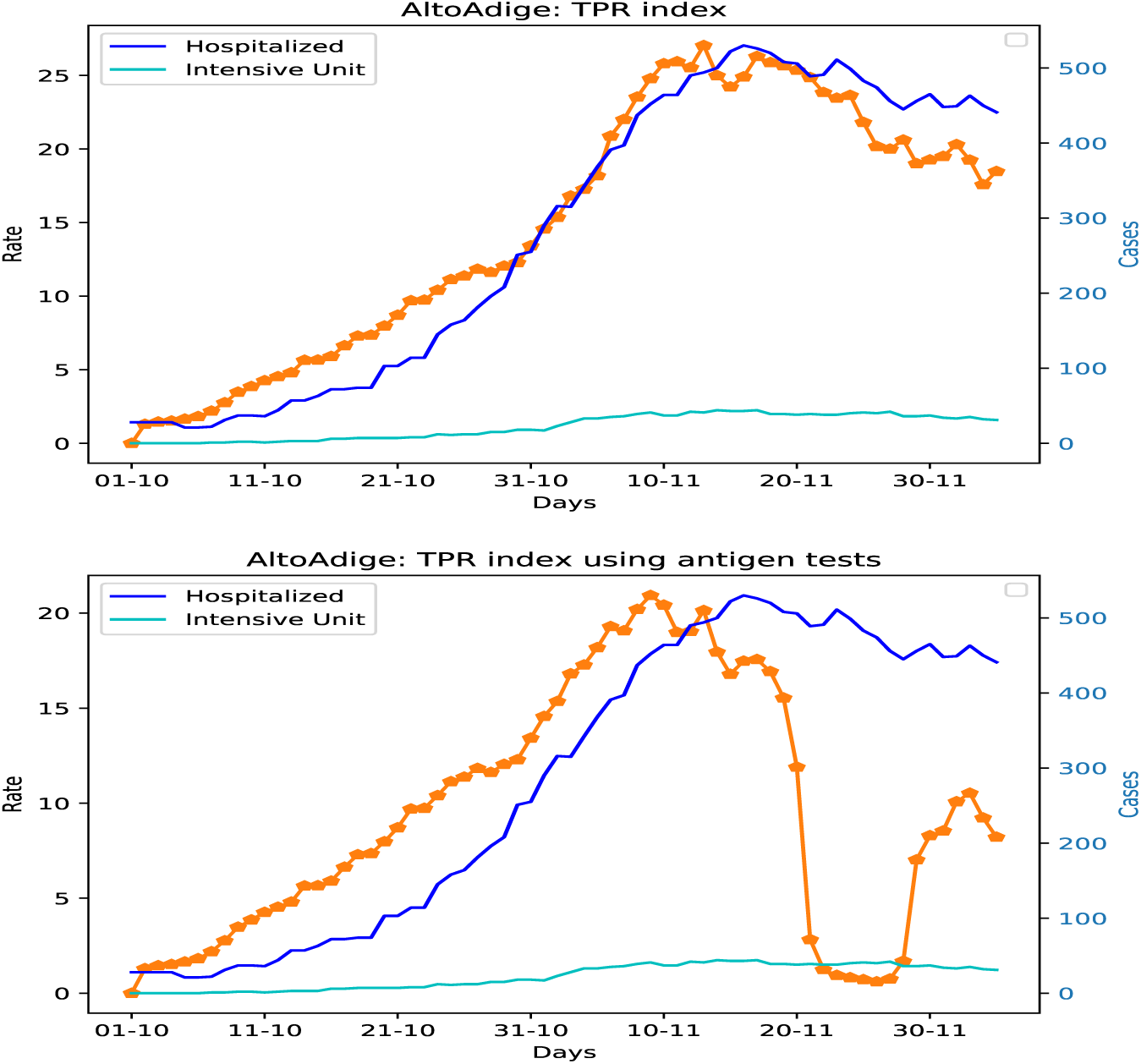
Alto Adige region: TPR index sliding compared with hospitalized data. The diagram in the bottom includes antigen tests in the estimation. The fast descent of the TPR index corresponds to the screening period from the 21th to the 23th of November 2020.

Considering that there are other regions, like Emilia Romagna that definitely have the same problem concerning availability of antigen tests data, we can argue that the TPR index maintains its forecasting capability in the second phase. Hence, it would be a recommendable tool to be used for decision support in these day, given that antigen tests data are now available.

## 5 Discussion

From the analysis of the course of the pandemic in the modelled regions several interesting considerations emerge.

### 5.1 The evolution of Veneto and Toscana regions

The course of the epidemic in Veneto was different from that of most other regions. Indeed the TPR index has not suddenly decreased after the peak of the 16th of November 2020, and after a peak of hospitalized people the 28th of November, the TPR index started to grow again slowly. A new TPR peak occurred the 9th of December, surpassing the previous one. This is the peak that was discovered later by ISS in the weekly report N. 31 of the 16th of December 2020 [15], concerning period between the 7th and 13th of December. Successively, 8 days later there was another peak of hospitalized people the 17th of December, but the growth rate of patients remained limited. In the last period, there was another TPR peak the 29th of December (see Figure 11), and after that another peak of hospitalized people the 4th of January. Once again, the growth percentage of hospitalized patients was not high, and successively the TPR began to fall steadily, and the number of patients in hospital started to lower. Given the consistent reduction of the TPR index, the number of patients will definitely decrease in the next few weeks, and this is effectively what happened.

**Figure 11:**
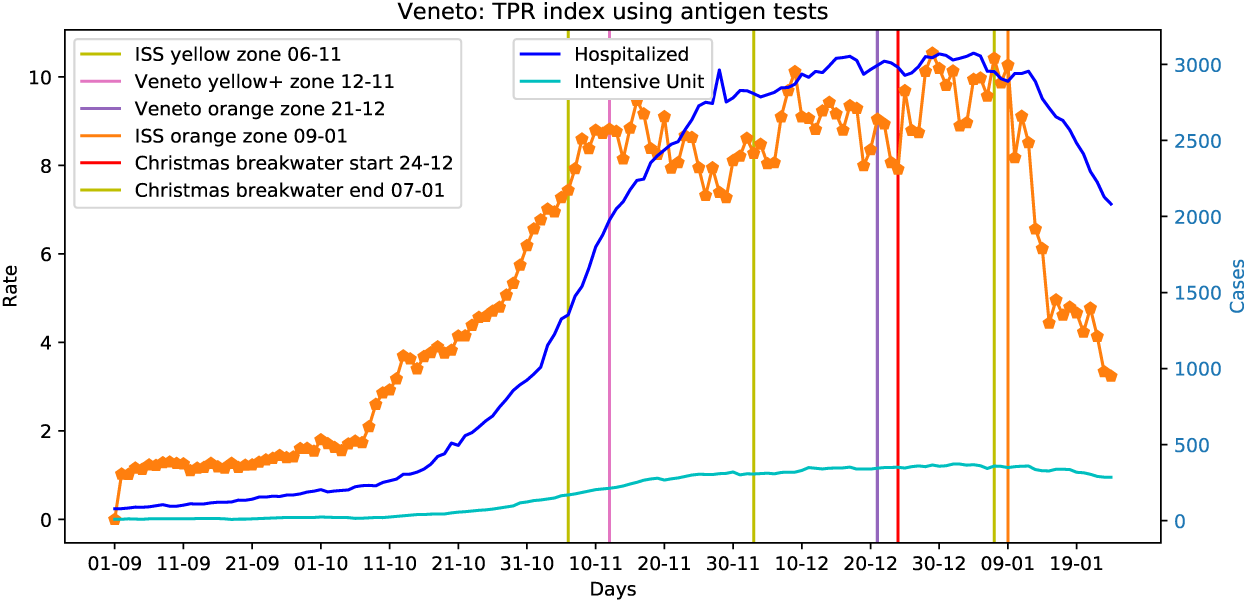
Veneto region: evolution of the TPR peaks and hospitalized peaks.

In summary, despite restrictions in Veneto were not so strong as those of other regions, the strategy characterized by a large number of tests and thorough tracking of contacts, is contending for the spread of the virus and prevented the collapse of the health care system, without excessively penalising the economy. The Toscana region is characterized by a more simple evolution of the TPR index curve, after the peak TPR dropped quickly, while it has started to rise gradually before Christmas, and the number of patients in hospitals has decreased more slowly, see Figure 12. Possibly, the different course of the epidemic in Toscana and Veneto is related to the adopted measures, we address this issue as the last point of this discussion.

**Figure 12:**
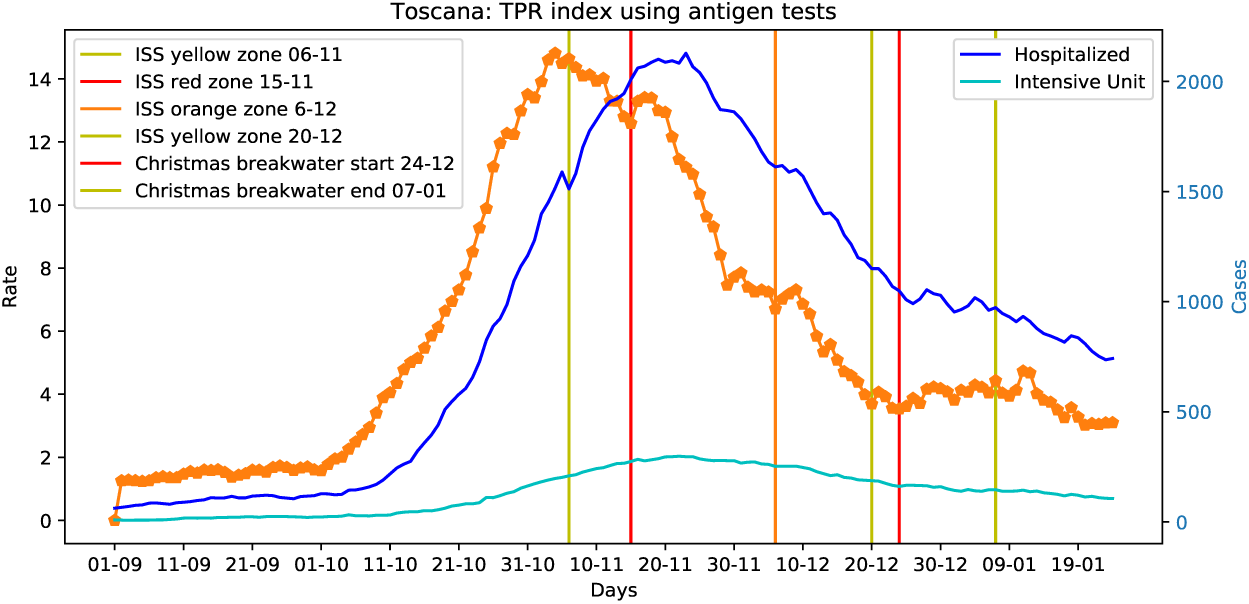
TOSCANA region: TPR index fall after the Christmass breakwater.

### 5.2 The impact of antigen tests on the TPR index

Another important issue which has been raised by some scientists, concern antigen tests sensitivity. Given that we assumed that positive antigen tests are in general confirmed by molecular ones, false positives do not pose a significant problem for the model.

Although, the false negatives percentages reported in the literature for novel antigen tests are not high [3, 4, 5], some scientists still doubt about their sensitivity in specific situations, for example with asymptomatic individuals [7], albeit the presented results are about salivary tests [6] not NS. However, the results of these studies strongly depend on the tests used, and actually there are many different tests around, see for example [2, 26]. It is not our intention here to shed light on this dispute, anyway FDA still recommend that it is not necessary to perform confirmatory molecular tests on individuals with negative antigen test results, if they are obtained during routine screening or surveillance [9], and they can actually be used for implementing cost effective diagnostic testing strategies [24].

Therefore, we focus on answering the question: may antigen tests failures have a significant impact on the TPR index trend? In other words, assuming that a significant percentage of antigen tests fail, will have an effect in the dates in which the peaks occur? We cut down the number of antigen tests by 15% and 30% (a reasonable upper bound for failures), and we computed the TPR index for the studied regions. What happened is that the TPR trend has remained unchanged without altering the dates of the peaks in Table 4. For example, figure 13 shows the TPR variations for Veneto, a small increment of the index is observed, but the trend does not changes. Hence, there is no evidence that unreliable antigen tests caused the increase of infections in Veneto.

**Figure 13:**
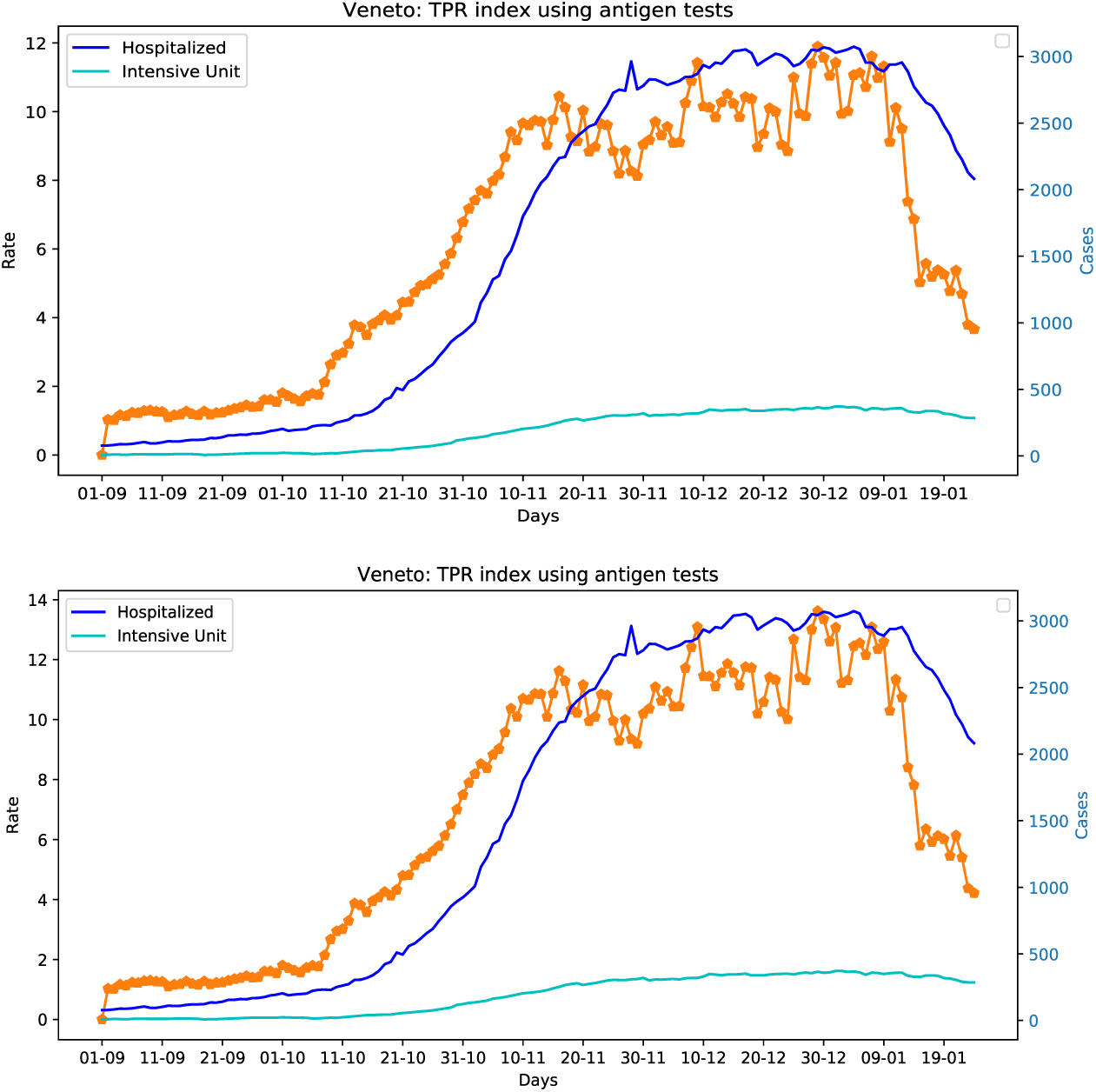
Veneto region: How the TPR index changes if we reduce the weight of antigent tests by 15% in the top or by 30% in the bottom. The trend and the peaks remains at the same with respect to Figure 11, we can argue that claiming that antigen tests caused the increase in infections is clearly unfounded.

### 5.3 The mass screening campaign of Alto Adige region

Considering the Alto Adige region, it is worth to report the effects of the mass screening on the TPR index and on hospitalized people which is illustrated in Figure 14. After the screening there were a considerable reduction of the TPR index (the lowest value in all Italy) and of the number of patients in hospitals. However, after one week the TPR index has begun to grow again. A factor that may had an important effect on the results of the screening campaign was the large number of exposed individuals. Indeed, if more than 3000 people were detected as positive in 3 days only, there could be at least the same number of people still incubating the virus, that was not possible to detect in the screening. This issue is about the course of the disease, and it is not related to possible antigen tests failures. Another observation concern the fact that the hospitalized patients peak was only 7 days after the TPR peak, possibly this depends from the fact that Alto Adige started to use antigen tests before the 6th of November, but these tests were not reported in official data. This hypothesis is confirmed by this study [7] which uses antigen tests, it was done in Alto Adige in the summer. In the last week of December an additional information was added to the provincia of Bolzano reports: the number of positive antigen tests. When they appeared, at the end of December, they were not included in the official data as new positive cases. However, given that when the new information was added (the 15th of January 2021), they were added to official positive cases, we updated the dataset using the same logic. In other words, we added the number of positive antigen tests reported by provincia of Bolzano to the number of new positive cases.

**Figure 14:**
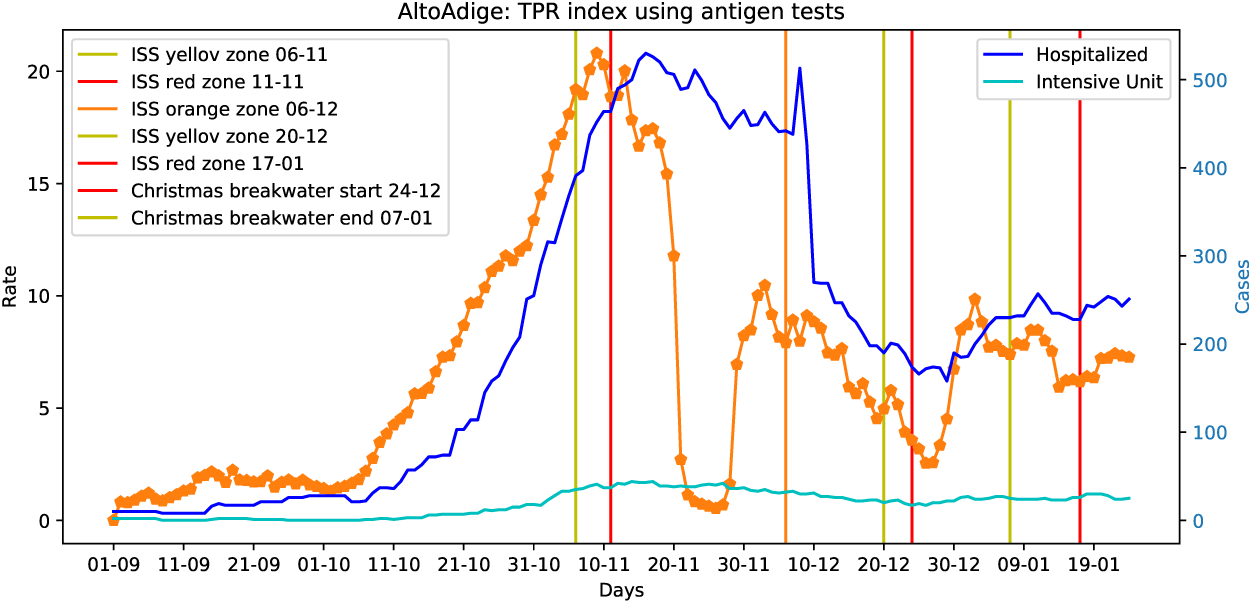
ALTOADIGE region: the effects of the November 2020 screening campaign.

### 5.4 Analysing Piemonte region

A further case that is worth to report concern the Piemonte region. In the middle of the month of December there was an evident break in the molecular tests series, when two hundred thousand tests were removed in one day, they were the antigen tests done starting from the 22th of October. As a result of this operation the TPR shot up through the roof, Figure 15 illustrates this effect. Thanks to the data made available in the web site of the region Piemonte health assessor [27], we were able to rebuilt the complete molecular tests series removing antigen tests (Figure 15 in the middle), and successively we introduced antigen tests as a separate series. With this dataset we were able to compute the TPR index for Piemonte. The results are that the relationship between the TPR index and hospitalized patients data can be also observed in Piemonte, and the ability to foresee peaks of COVID-19 patients admitted in hospital was restored (Figure 15 in the bottom).

**Figure 15:**
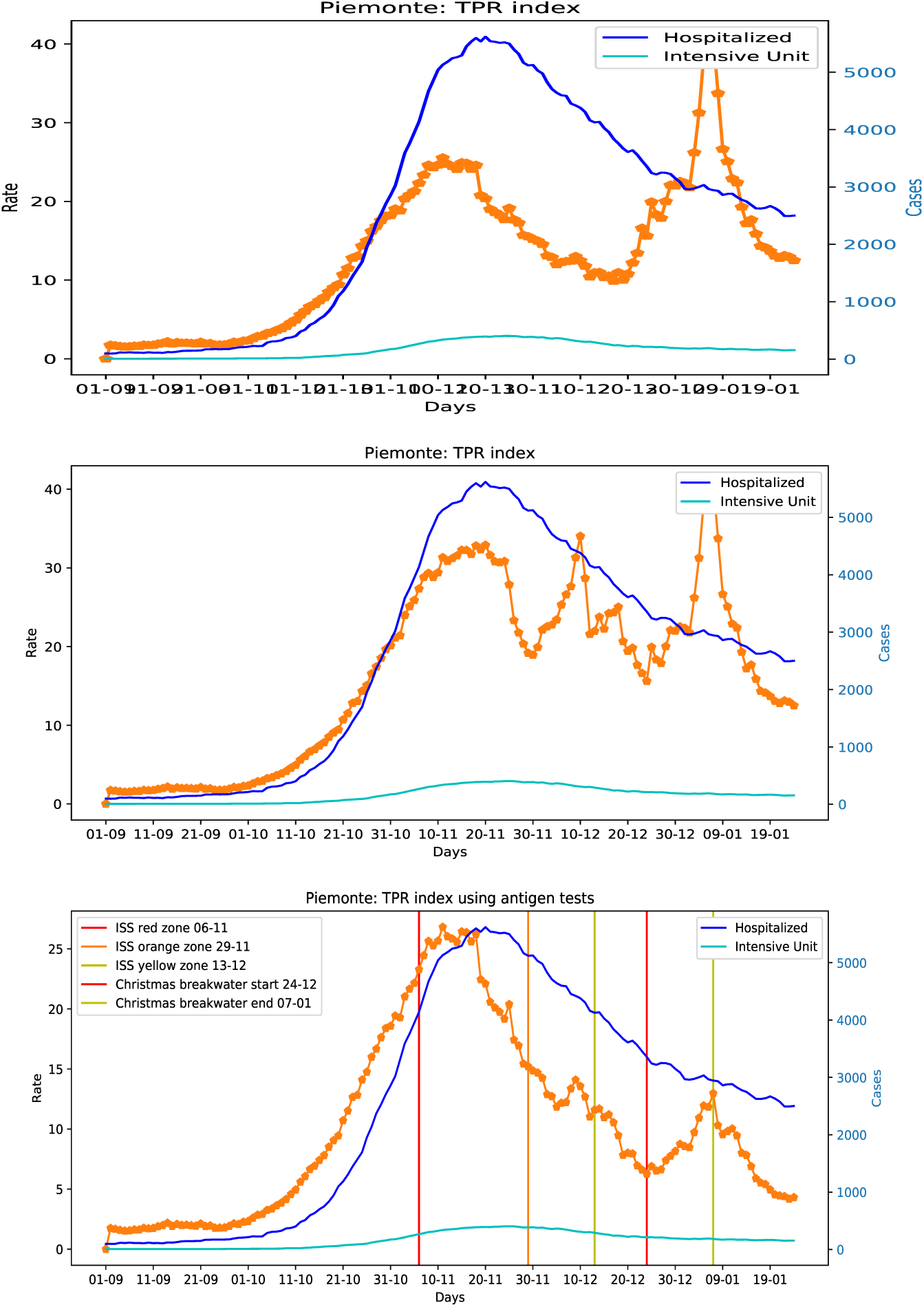
PIEMONTE region TPR index trend: in the top the broken series; in the middle the series removing antigen tests; in the bottom the series using antigen tests.

### 5.5 TPR index trend vs control measures

The last point of this discussion concerns the relationship between the TPR index and the Italian minister’s of health three-tiered system issued at the beginning of November 2020 to combat the spread of COVID-19. In this system regions are classified in three zones with increasing restrictions on the basis of ISS reports [13, 14, 15].

- *Yellow zone:* moderate risk, bars and restaurants must stop service at 18:00; cinemas, theatres, swimming pools and gyms are closed. People are not allowed to be out of their home from 22:00 to 05:00, except for work or health reasons.
- *Orange zone:* medium-high risk, most non-essential shops can stay open, while bars and restaurants must close, apart from for takeaway services; people can move freely within their cities, but cannot travel elsewhere.
- *Red zone:* high-contagion-risk, non-essential shops and markets are closed and residents are only allowed to leave their homes for work, health reasons or emergencies.

Additionally, region Veneto has decreed strengthened yellow and orange zones with closure of shopping malls at weekends, and banning the movements from home towns after 14.00.

Figures 11, 14, 12, and 15 illustrate the relationship between the course of TPR index and the measures adopted for Italian regions. Several considerations can be done analysing these figures.

The first observation that can be made concerns the possibility of anticipating pandemic control interventions, if the continuous and significant increasing of TPR index had been observed at the beginning of October 2020. This effect was clear for all the 4 analysed regions.

Another, issue concern yellow zones, it appears that they have not a significant impact on reducing the TPR index, while both orange and red zones effectively do it.

Finally, for all the considered regions the effect of the Christmas partial lock down has led to a lowering of the TPR index in the successive weeks. We define this phenomena *breakwater* effect, as in the case of sea waves a sequence of bulwarks has a significant cost effective impact on the reduction of effects of stormy seas, a sequence of short lock down (3/4 days) seems to be able to reduce the impact of the COVID-19 on the population. Moreover, we noticed that the reduction of the TPR index was greater if the higher the number of tests done in the same period. Veneto did the higher number of tests, Piemonte, Toscana and Alto Adige followed in order.

## 6 Standardizing antigen tests data

The simple TPR estimation method we exploited works fine, even if the information we have is limited to the number of antigen tests done in a given day only.

Anyway, the precision of this index could be improved, if more structured information will be made available, eventually including data on vaccinate individuals. Indeed, considering that COVID-19 diagnosis using antigen tests only are also possible (see the report [16]), additional information is needed to represent this eventuality. More precisely, the data that should be collected for each day are the following:

- *dayA*: the number of antigent tests;
- *dayAp*: the number of positive antigen tests;
- *dayAd* : the number of COVID-19 diagnosis based only on antigen tests.
- *dayV* : the number of vaccined persons.

The new factor Φ^*V*^ will be:

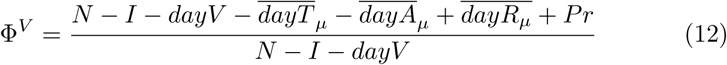

where *Pr* is the number of repeated tests, computed as follows:

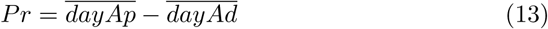

if the *dayAp* data are not made available, they can still be computed using the approach presented above (Equation 7). The new TPR index 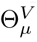 which includes antigen tests and vaccinate individuals is defined by the following formula:

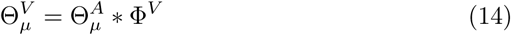

which would allow to estimate the TPR in a context in which the number of susceptible continuously decreases.

Although, the data on antigen tests made recently available in the Protezione Civile site are slightly different, they can be easily used to compute the proposed index. They include, the number of molecular tests, the number of antigen tests, the number of positive molecular tests, and the number of COVID-19 diagnosis based only on antigen tests *dayAd*. Thus *dayAp*, the number of positive antigen tests is not provided, while it can still be estimated using the proposed method.

## 7 Conclusion

We have shown that a well defined daily test positivity rate is a powerful index that can be used to control the evolution of the COVID-19 pandemic. Indeed, in some situations it would be possible to forecast peaks of hospitalized people and the associated variations in percentage. Antigen tests data have to be considered for making TPR reliable. The behaviour of the proposed index is studied using the data of Italian regions, considering both the first and the second phase of the pandemic. We extended the Italian Protezione Civile dataset [17] with antigen tests for Alto Adige, Toscana Veneto and Piemonte. This extended dataset allowed us to study the impact of antigen tests in epidemic control strategies and the effects of screening campaigns on the population.

From this study two promising response strategies emerge for COVID-19: adopting early interventions if the TPR index starts to grow significantly for more then 10-15 days; a *breakwater* cost effective strategy done of short lock down (3/4 days a week) repeated for two or three weeks, combined with an extensive testing campaign.

To conclude, a extended TPR index including a more complete formalization of antigen tests and the number of vaccinate people is proposed. Future work will concern the definition of rules of thumb to indicate measures to adopt if certain TPR patterns are discovered. Although, the data on antigen tests are now available in the Protezione Civile web site [17], a further effort should be done to reconstruct past time series for all the Italian regions. This would allow scientists to study in a more accurate way the evolution of the TPR index combined with the adopted measures and the effects of the proposed strategies and screening policies on the pandemic course.

## Data Availability

The used data are available in the following sites:

https://github.com/pcm-dpc

http://www.cs.unibo.it/∼gaspari/www/myreg.csv

## Acknowledgements

The author would like to thank the Italian Civil Protection Department, and all the staff involved for providing the data of the outbreak used in this study. Also, the author would like to thank the Bolzano province (Alto Adige) for providing the data of the screening campaign, and for their daily comprehensive coronavirus reports including antigen tests. Finally, the author would like to thank the Veneto region for the high technical quality and communication skill of the team involved in the management of the outbreak. The idea of standardizing data of existing COVID-19 tests was given by Dr. Francesca Russo, Regione Veneto.

## Availability of data and material

The used data are available in this web site: https://github.com/pcm-dpc.

The extended dataset also including antigen tests data of veneto, Toscana, Alto Adige and Piemonte is available here: http://www.cs.unibo.it/~gaspari/www/myreg.csv

## Code availability

The code is available upon request.

## Footnotes

1 The day in which the first positive individual was detected in a given region

2 The positivity to SARS-CoV-2 infection was followed by home isolation measures without carrying out a confirmative PCR test [23]

